# Cellular and humoral immunogenicity of a SARS-CoV-2 mRNA vaccine in patients on hemodialysis

**DOI:** 10.1101/2021.05.26.21257860

**Authors:** Monika Strengert, Matthias Becker, Gema Morilla Ramos, Alex Dulovic, Jens Gruber, Jennifer Juengling, Karsten Lürken, Andrea Beigel, Eike Wrenger, Gerhard Lonnemann, Anne Cossmann, Metodi V. Stankov, Alexandra Dopfer-Jablonka, Philipp D. Kaiser, Bjoern Traenkle, Ulrich Rothbauer, Gérard Krause, Nicole Schneiderhan-Marra, Georg M.N. Behrens

## Abstract

**Background:** Patients with chronic renal insufficiency on intermittent hemodialysis face an increased risk of COVID-19 induced mortality and impaired vaccine responses. To date, only few studies addressed SARS-CoV-2 vaccine elicited immunity in this immunocompromised population.

**Methods:** We assessed immunogenicity of the mRNA vaccine BNT162b2 in at risk dialysis patients and characterized systemic cellular and humoral immune responses in serum and saliva using interferon γ release assay and multiplex-based cytokine and immunoglobulin measurements. We further compared binding capacity and neutralization efficacy of vaccination-induced immunoglobulins against emerging SARS-CoV-2 variants of concern B.1.1.7, B.1.351, B.1.429 and Cluster 5 by ACE2-RBD competition assay.

**Findings:** Patients on intermittent hemodialysis exhibit detectable but variable cellular and humoral immune responses against SARS-CoV-2 and variants of concern after a two-dose regimen of BNT162b2. Although vaccination-induced immunoglobulins were detectable in saliva and plasma, both anti-SARS-CoV-2 IgG and neutralization efficacy was reduced compared to controls. Similarly, T-cell mediated interferon γ release after stimulation with SARS-CoV-2 spike peptides was significantly diminished.

**Interpretation:** Quantifiable humoral and cellular immune responses after BNT162b2 vaccination in individuals on intermittent dialysis are encouraging, but urge for longitudinal follow-up to assess longevity of immunity. Diminished virus neutralization and interferon γ responses in face of emerging variants of concern may favor this at risk population for re-vaccination using modified vaccines at the earliest opportunity.

**Funding:** Initiative and Networking Fund of the Helmholtz Association of German Research Centers, EU Horizon 2020 research and innovation program, State Ministry of Baden-Württemberg for Economic Affairs, Labor and Tourism.

**Research in the context:** *Evidence before this study:* Patients on dialysis tend to have a reduced immune response to both infection and vaccination. We searched PubMed and MedRxiv for studies including search terms such as “COVID-19”, “vaccine”, and “dialysis” but no peer-reviewed studies to date assessed both SARS-CoV-2 specific B- and T-cell responses, mucosal immunoglobulins, and considered the impact of SARS-CoV-2 variants of concern in this at risk population.

*Added value of the study:* We provide a comprehensive functional characterization of both T- and B-cell responses following a two-dose regimen of BNT162b2 in at risk patients on maintenance hemodialysis. More importantly, to the best of our knowledge, we assess for the first time binding and neutralization capacity of vaccination-induced circulation and mucosal antibodies towards emerging SARS-CoV-2 variants of concern in an immunocompromised population.

*Implications of all the available evidence:* Patients on maintenance hemodialysis develop a substantial cellular and humoral immune response following the BNT162b2 vaccine. These findings should encourage patients on intermittent hemodialysis to receive the vaccine. However, we suggest continuing additional protection measures against variants of concern in this at risk population until longevity of the vaccine response is fully evaluated.

## 1. Introduction

Since its emergence in late 2019, SARS-CoV-2 has become a global pandemic with more than 166 million confirmed cases and 3·46 million deaths (as of 24.05.21) (1). Vulnerable populations such as the elderly, immunocompromised or those suffering from chronic conditions or requiring continual medical intervention such as dialysis are at risk of severe COVID-19 disease and associated death (2).

Although a series of vaccines have been developed, tested and approved at unprecedented speed, only one vaccine study for NVX CoV2373 has enrolled patients with chronic diseases such as chronic kidney disease to assess efficacy and safety of SARS-CoV-2 vaccination within this vulnerable population (3). Patients on maintenance dialysis are a particularly high-risk group, as renal disease has been identified as a key risk factor for severe COVID-19 (4-7), while at the same time their regular need for therapy does not allow them to self-isolate and reduce contacts to avoid infection. A recent study also suggested that seroreversion following natural SARS-CoV-2 infection is faster in dialysis patients compared to the general population further increasing the risk of re-infection (8). Other studies have also identified that vaccine efficacy is reduced in dialysis patients, as seen by the attenuation of antibody titers following vaccination with SARS-CoV-2 vaccines (9-14). Additional data on the efficiency of SARS-CoV-2 vaccination is urgently needed not only for risk mitigation but also to assess whether additional protective measures during therapy must be put in place as seen by the impaired infection and vaccination-induced responses for influenza A and hepatitis B (15-17). While antibody titers have already been characterized within vaccinated dialysis patients, little is known about their neutralization potential, particularly in light of the increasingly appearing SARS-CoV-2 variants of concern (VoC) which threaten the success of vaccination programs (18).

To assess efficacy of SARS-CoV-2 vaccination in dialysis patients, we characterized cellular and humoral immune responses in serum and saliva of dialyzed and non-dialyzed individuals after vaccination with the mRNA vaccine BNT162b2. Considering the increasing presence of mutated SARS-CoV-2 strains, we further compared binding capacity and neutralization efficacy of vaccination-induced immunoglobulins against emerging variants of concern (VoC) such as B.1.1.7, B.1.351, B.1.429 and Cluster 5.

## 2. Methods

### 2.1 Study design and sample collection

Following written informed consent, heparinized blood samples from hemodialysis patients were taken before start of dialysis using the vascular access which was either an arterio-venous fistula or a central venous catheter or by venipuncture from health care workers (control). Details about the study population are listed in Table 1, Table S2 and S3. One hemodialysis patient was previously tested positive for SARS-CoV-2 by routine PCR screening. As subsequent antibody testing was negative for SARS-CoV-2 IgG, this patient received BNT162b2 and was not excluded from our study. The standard two-dose regimen of Pfizer was given 21 days apart, followed by blood collection for analysis 21 days after the second dose. Plasma was obtained from lithium heparin blood (S-Monovette, Sarstedt, Germany) and stored at -80°C until use. Whole blood samples were used immediately for interferon γ release assay (IGRA). For saliva collection, all individuals spat directly into a collection tube. To inactive saliva samples, Tri(n-butyl) phosphate (TnBP) and Triton X-100 were added to final concentrations of 0·3% and 1%, respectively (19). Saliva samples were then frozen at -80°C until further use.

**Table 1.** Characteristics of vaccinated study participants **-** IQR - Inter Quartile Range. BMI - Body Mass Index.

| Characteristics | Non-dialysis control group (n=34) | Hemodialysis group (n=81) |
| --- | --- | --- |
| Age (years), median (IQR) | 54·5 (45-60) | 69 (60-78) |
| Gender (female, n, %) | 28 (82·35) | 34 (41·98) |
| Days since start of hemodialysis (median, IQR) | n. a | 1371 (638-2302) |
| Immunosuppressive medication (n, %) | 0 (0) | 10 (12·34) |
| Co-morbidities |  |  |
| Obesity (BMI, >30) | 8 (23·53) (1 NA) | 18 (22·22) |
| Diabetes mellitus (n, %) | 1(2·94) | 22 (27·16) |
| Cardiovascular disease (n, %) | 0 (0) | 39 (48·15) |

### 2.2 Ethics statement

The study was approved by the Internal Review Board of Hannover Medical School (MHH, approval number 8973_BO-K_2020).

### 2.3 Bead coupling

Coupling of antigens to spectrally distinct MagPlex beads was done by EDC/s-NHS coupling for all standard MULTICOV-AB antigens (20). RBDs from variants of concern were coupled using Anteo coupling (#A-LMPAKMM-10, Anteo Tech Reagents, Australia) following the manufacturer’s instructions (21).

### 2.4 MULTICOV-AB

Antibody titers and binding was analyzed using MULTICOV-AB, a multiplex immunoassay which simultaneously analyses 20 antigens, as previously described (20). The full list of antigens included in this study can be found in Table S1. Plasma samples were diluted 1:400 while saliva samples were diluted 1:12 (21). Briefly, antigens were immobilized on spectrally distinct populations of MagPlex beads (as above) and combined into a single bead mix. Samples were combined with the bead mix, incubated for 2 h at 21°C and then washed using a microplate washer to remove unbound antibodies. Bound antibodies were detected following a 45 min incubation at 21°C with R-phycoerythrin labeled goat-anti-human IgG (Jackson ImmunoResearch Labs, United Kingdom, Cat# 109-116-098, Lot# 148837, RRID: AB_2337678, used at 3 µg/mL) or IgA (Jackson ImmunoResearch Labs, Cat# 109-115-011, Lot# 143454, RRID: AB_2337674, used at 5 µg/mL) as secondary antibodies. Following another washing step, beads were re-suspended and then measured using a FLEXMAP3D instrument (Luminex, Texas, US) using the following settings: Timeout 80 sec, Gate: 7500-15000, Reporter Gain: Standard PMT, 40 events. Each sample was measured once. As a control, three quality control (QC) samples were included on each plate. Raw median fluorescence intensity (MFI) values or normalized values (MFI/MFI of QC samples (21)) are reported.

### 2.5 ACE2-RBD competition assay

To determine neutralization, an ACE2-RBD competition assay was carried out as previously described (21) Briefly, biotinylated ACE2 was added to the assay buffer to final concentration of 500 ng/mL for all samples. Samples were then mixed with MULTICOV-AB bead mix (see above) and incubated for 2 h at 21°C, 750 rpm. After washing, ACE2 was detected using Streptavidin-PE (2 µg/mL, #SAPE-001, Moss, Maryland, US) by incubating the sample for 45 min at 750rpm. After an additional wash step and resuspension, samples were measured on a FLEXMAP3D instrument (same settings as MULTICOV-AB). As control, 500 ng/mL ACE2 was used. For analysis, MFI values were normalized against the control wells. All samples were measured once.

### 2.6 Euroimmun ELISA QuantiVac

To further validate plasma IgG levels measured by MULTICOV-AB, samples were further analyzed using the Anti-SARS-CoV-2-QuantiVac-ELISA IgG (Euroimmun, Germany) according to the manufacturer’s instructions. Plasma samples were diluted 1:400 to achieve assay linearity.

### 2.7 Interferon γ release assay (IGRA)

SARS-CoV-2-specific T-cell responses were determined by measuring IFNγ production upon SARS-CoV-2 antigen stimulation using the SARS-CoV-2 Interferon Gamma Release Assay IGRA (Euroimmun, Germany). Briefly, 0·5 mL full blood were stimulated with peptides of the SARS-CoV-2 S1 domain of the Spike protein for a period of 20-24 h. Negative and positive controls were carried out according to the manufacturer’s instruction. Following stimulation, supernatants were isolated through centrifugation and IFNγ measured using ELISA. The remaining supernatant was stored at -80°C. Background signals from negative controls were subtracted and final results calculated in mIU/mL using standard curves. IFNγ concentrations >200 mIU/mL were considered as reactive, the upper limit of reactivity was 2000 mIU/mL.

### 2.8 Cytokine measurements

Supernatants of SARS-CoV-2 antigen stimulated blood cells were prepared and isolated as explained for the detection of IFN-γ by SARS-CoV-2 Interferon Gamma Release Assay (IGRA) and analyzed by LEGENDplex™ using the Human Essential Immune Response Panel (Bio Legend, California, US) for L-4, IL-2, CXCL-10 (IP-10), IL-1β, TNFα, CCL-2 (MCP-1), IL-17A, IL-6, IL-10, IFNγ, IL-12p70, CXCL-8 (IL-8), TGFβ1) according to manufacturer’s instructions. Analysis was performed using an LSR II flow cytometer (Becton Dickinson, Germany) and data analyzed using the LEGENDplex™ Data Analysis Software Suite.

### 2.9 Data Analysis

RStudio (Version 1.2.5001), with R (version 3.6.1) was used for data analysis and figure generation. The additional packages “beeswarm” and “RcolorBrewer” were used only for data depiction purposes. The type of statistical analysis performed (when appropriate) is listed in the figure legends. Figures were exported from Rstudio and then edited using Inkscape (Inkscape 0.92.4). Spearman’s ρ coefficient was calculated in order to determine correlation between IGRA results and antibody responses or neutralization using the “cor” function from R’s “stats” library. Mann-Whitney U test was used to determine difference between signal distributions between dialyzed and control groups using the “wilcox.test” function from R’s “stats” library. Pre-processing of data such as matching sample metadata and collecting results from multiple assay platforms was performed in Excel 2016.

## 3. Results

### 3.1 Dialyzed patients have reduced antibody titers following vaccination

To characterize the vaccination response in patients on maintenance dialysis, we measured IgG and IgA levels in plasma 21 days after the second dose of Pfizer BNT162b2 using MULTICOV-AB, a multiplex immunoassay containing antigens from Spike and Nucleocapsid proteins of both SARS-CoV-2 and the endemic human coronaviruses (hCoVs) (20). As a control group, 34 samples from healthcare workers vaccinated at the same time points as the 81 patients on hemodialysis were used. (Detailed information on the study population can be found in Table 1, Table S2 and S3.) As indicated by the lack of a significant anti-Nucleocapsid (N) IgG or IgA response, none of the study participants had been previously infected or seroconverted after a SARS-CoV-2 infection (data not shown). IgG responses towards the original B.1 isolate in vaccinated dialysis patients were significantly reduced (p<0·0001) and more variable (Fig. 1a.) than in the control group, which reached the upper limit of detection of the assay, as seen previously (21). Interestingly, plasma IgA responses in the dialysis group were comparable to the control group (Fig. 1b, p=0·38). Overall, within our dialysis population, four from 81 vaccinated individuals (4·92%) were classified as serologic non-responders with antibody titers below the cut-off. As an additional control, S1 IgG titers were measured using a commercial assay (Fig. S2), which identified the same pattern of a significantly diminished antibody response in dialyzed patients (272·3 RU/mL) compared to non-dialyzed individuals (456·8 RU/mL, p<0·0001). As SARS-CoV-2 is a mucosal-targeted virus, we also collected saliva from our vaccination cohort and assessed IgG and IgA levels using MULTICOV-AB. When examining antibody titers found in saliva, dialyzed individuals had significantly lower IgG titers (p=0·0007) but similar IgA titers (p=0·70) to the control group (Fig. 1c and d). To examine responses towards emerging variants of concern (VoC), Spike-receptor binding domains (RBD) of the B.1.1.7 (UK), B.1.351 (South African), Cluster 5 (Mink) and B.1.429 (LA) variants were included as part of MULTICOV-AB (21). As expected (18, 21), antibody binding towards B.1.1.7 in both dialyzed and non-dialyzed individuals was comparable to B.1 (original isolate), while binding was clearly reduced for B.1.351 (Fig. 1a). Antibody binding for Cluster 5 and B.1.429 was similar to the B.1 isolate for both groups (Fig. S1). As part of the MULTICOV-AB antigen panel, we also analyzed the humoral response towards endemic CoV S1 and N protein, but found no general significant difference between control group and dialysis patients (Fig. S3).

**Fig. 1.**
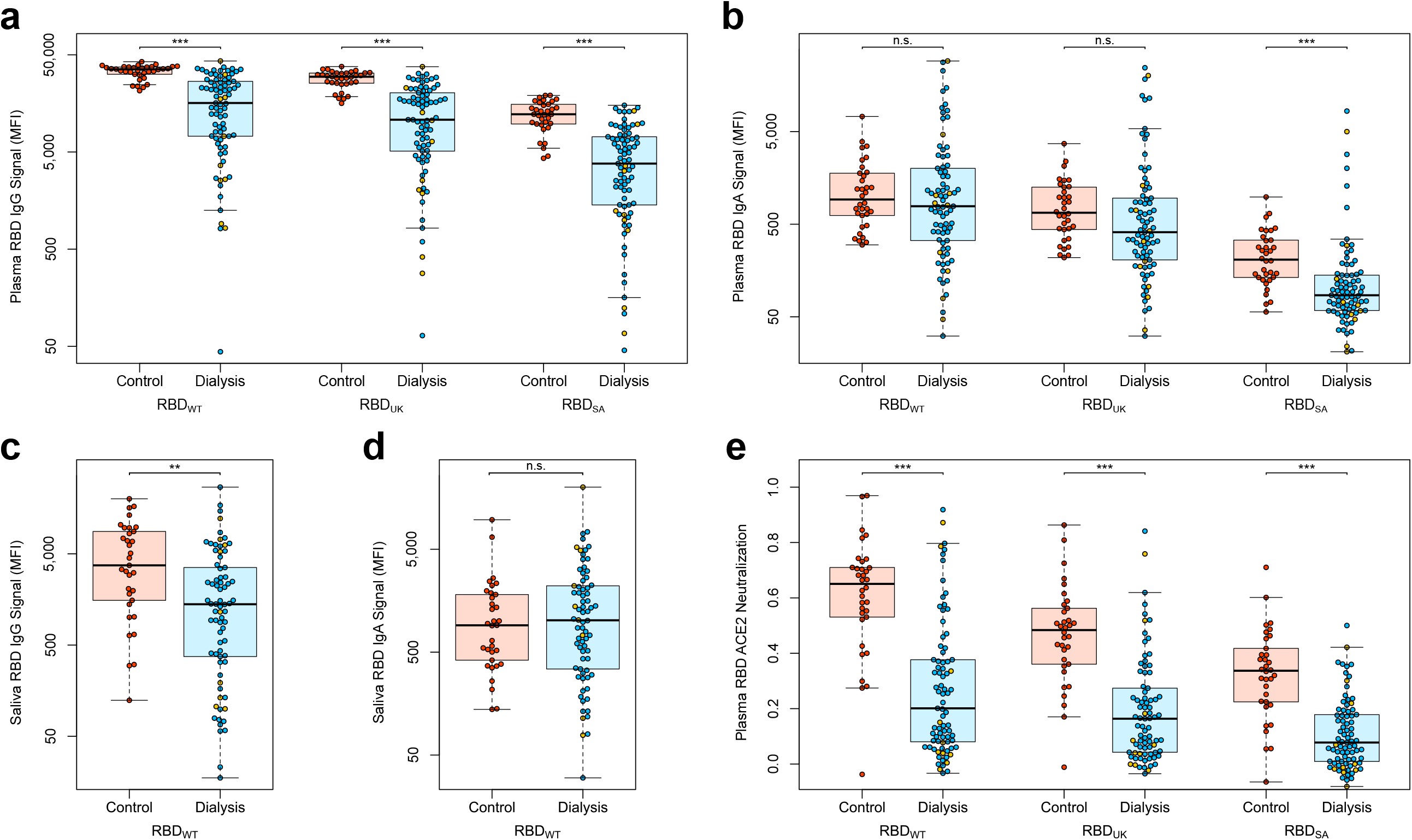
Humoral immune response in hemodialyzed individuals after vaccination with Pfizer BNT162b2. IgG (a, c), IgA response (b, d) and neutralizing capacity of IgG (e) towards the indicated SARS-CoV-2 WT (B.1), UK (B.1.1.7), South African (B.1.351) in plasma (a, b, e) or saliva (c, d) from controls (red circles, n=34), individuals on maintenance dialysis (blue circles, n=71) and hemodialyzed individuals on immunosuppressive medication (yellow circles, n=10) 21 days post second vaccination was measured using MULTICOV-AB (a, b, c, d) or an ACE2-RBD competition assay (e). Data is displayed as median fluorescence intensity (MFI) (a-d). Neutralization capacity is displayed as ratio where 1 indicates maximum neutralization and 0 no neutralization (e). Saliva (c, d) was collected from 33 controls (red circles), 65 individuals on maintenance dialysis (blue circles) and from 9 individuals on maintenance dialysis and immunosuppressive medication (yellow circles). Boxes represent the median, 25th and 75th percentiles, whiskers show the largest and smallest non-outlier values. Outliers were determined by 1·5 times IQR. Statistical significance was calculated by Mann-Whitney-U (two-sided). Significance was defined as ^*^<0·01, ^**^<0·001, ^***^<0·0001 or n.s.>0·01.

### 3.2 Neutralization is reduced in dialysis patients after vaccination

To assess neutralizing potency of plasma towards both the original B.1 isolate and VoC RBDs, we used a previously described ACE2-RBD competition assay (21). Neutralization across both wild-type and all VoCs measured was significantly reduced in dialyzed individuals compared to non-dialyzed (all p<0·0001) (Fig. 1e, Fig. S4). As expected, there were differences between the variants of concern themselves, with B.1.351 having the lowest neutralization for both control and dialyzed individuals. However, responses were comparably low for all VoC tested for patients on intermittent dialysis and additional immunosuppressive medication in the dialysis group further reduced neutralizing potency with all samples located in the 25^th^ quartile (Fig. 1e).

### 3.3 T-cell response to SARS-CoV-2 vaccination is diminished in dialyzed individuals

As clinical studies have suggested that both cellular and humoral response can confer protection from a SARS-CoV-2 infection (22), we also assessed vaccination-induced T-cell responses by IFNγ release assay and characterized cytokine and chemokine responses after stimulation with a Spike S1-derived peptide pool by multiplex cytokine measurements. Consistent with reduced anti-Spike S1 IgG and anti-RBD IgG levels (Fig. 1a, Fig. S2), IGRA showed significantly lower levels of IFNγ released in the supernatants of stimulated T-cells from vaccinated patients on maintenance dialysis (p<0·0001, Fig. 2a). In addition, within the control group, all but one T-cell sample were classified as reactive by IGRA whereas only 71·6 % were within the hemodialyzed group. Of the 12 analyzed cytokines beyond IFNγ, only IL-8 and CCL-2 (both p<0·0001) were significantly different between the two immunized groups, whereas no other Th1 type cytokines such as TNFα or IL-2 accompanied the IFNγ response (Fig. 2b-f, Fig. S5).

**Fig. 2.**
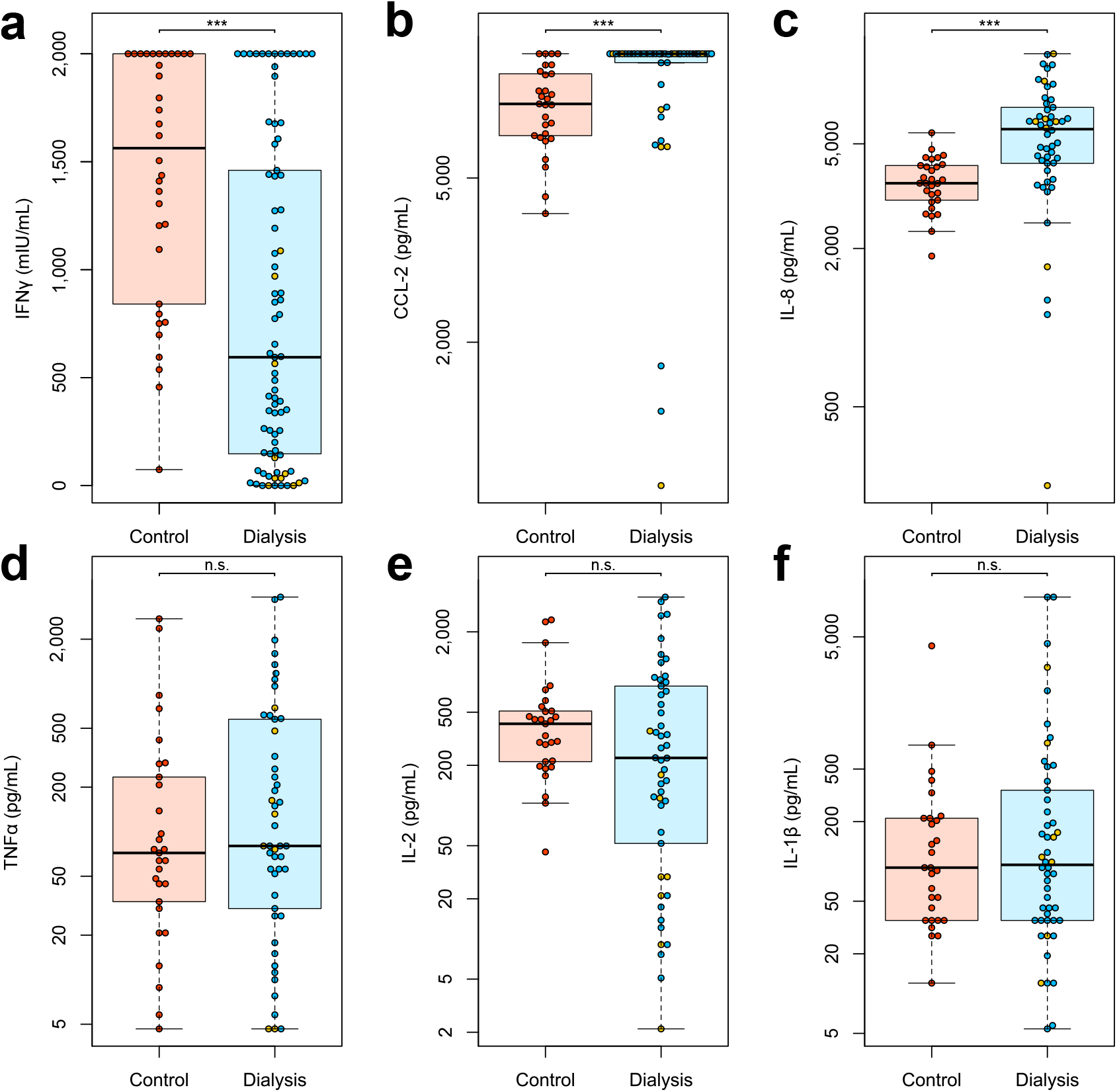
Cellular immune response in hemodialyzed individuals after vaccination with Pfizer BNT162b2. Whole blood from vaccinated controls (red circles, n=34), individuals on maintenance dialysis (blue circles, n=71) and hemodialyzed individuals on immunosuppressive medication (yellow circles, n=10) 21 days post second vaccination was *ex vivo* stimulated using a SARS-CoV-2 Spike S1 specific peptide pool. Supernatant fractions were analyzed by interferon γ release assay (IGRA, a) or bead-based multiplex-cytokine assay for CCL-2 (b), IL-8 (c), TNFα (d), IL-2 (e) and IL-1β (f). Data is shown in mIU/mL for IGRA or pg/mL for the multiplex-cytokine assay. T-cells were classified as reactive if IFNγ was >200 mIU/mL. IGRA (a) was carried out with samples from all study participants. Bead based-cytokine measurements (b-f) were performed with samples from 29 control, 42 hemodialyzed and 8 hemodialyzed individuals on immunosuppressive medication. Samples that were classified as above upper or below the lower limit of detection of the cytokine assay or the IGRA are shown at the respective limit. Boxes represent the median, 25th and 75th percentiles, whiskers show the largest and smallest non-outlier values. Outliers were determined by 1·5 times IQR. Statistical significance was calculated by Mann-Whitney-U (two-sided). Significance was defined as ^*^<0·01, ^**^<0·001, ^***^<0·0001 or n.s.>0·01.

As of now there are no defined correlates of protection against a SARS-CoV-2 infection and relative importance of cellular versus humoral response is equally undefined (23, 24), so we correlated B- and T-cell responses in our vaccination cohort. Comparable to other studies examining vaccination responses of BNT162b2 in a similar setting (25), we observed a moderate correlation between T-cell responses measured by IGRA and B-cell responses determined by RBD B.1-specific IgG levels (ρ=0·56, Fig. 3a) and RBD B.1 IgG-neutralizing potency (ρ=0·55, Fig. 3b) and a skew towards increased B-cell reactivity in both control group and individuals on hemodialysis.

**Fig. 3.**
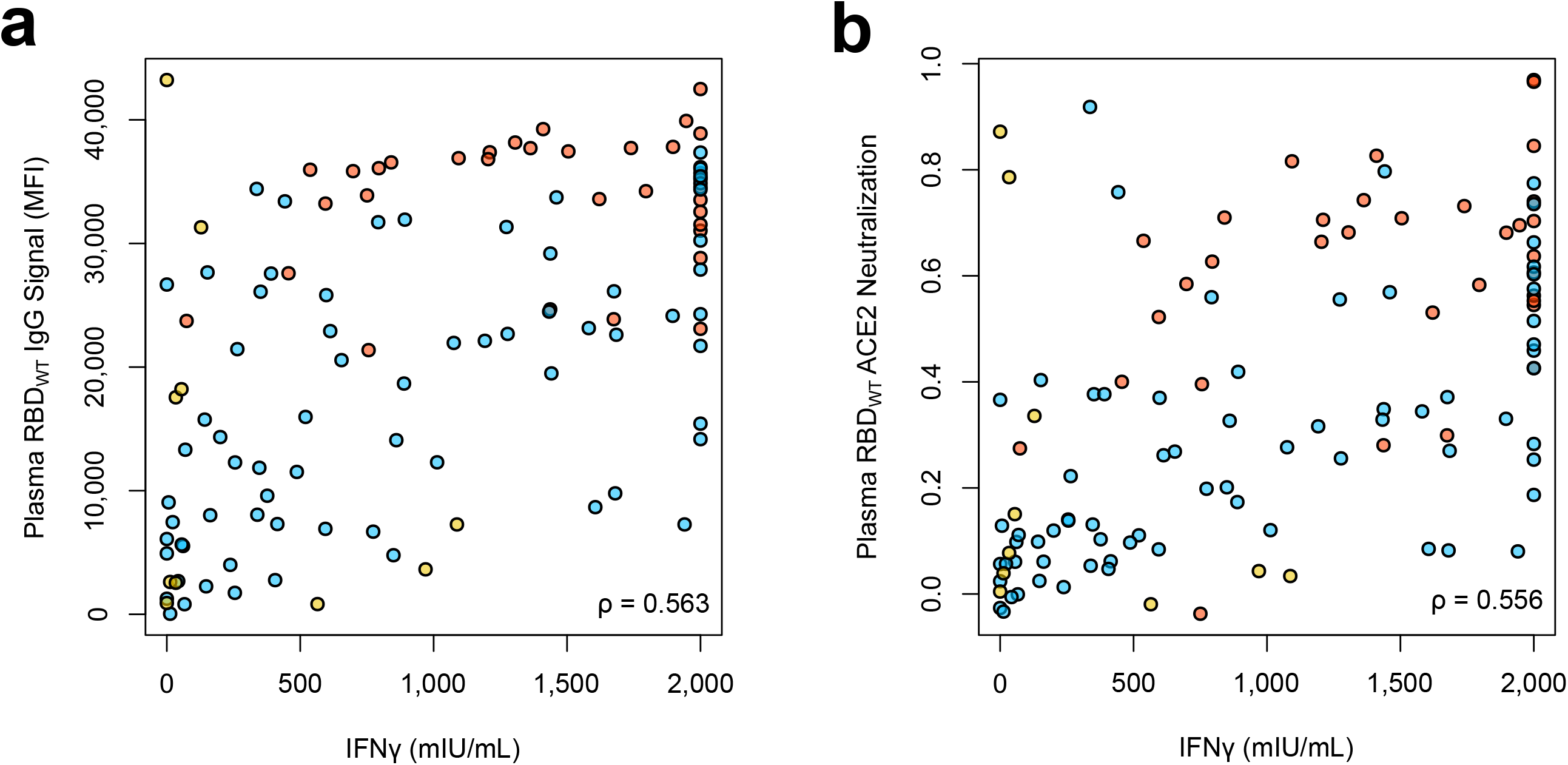
Relationship of cellular and humoral immune response after vaccination with Pfizer BNT162b2. T-cell responses assessed by IGRA for IFNγ (mIU/mL) and B-cell responses assessed by MULTICOV-AB IgG binding to WT RBD B.1 (a) or ACE2-WT RBD (B.1) competition assay (b) were plotted for correlation analysis in the vaccinated control group (red circles, n=34), in the vaccinated hemodialyzed group (blue circles, n=71) and in hemodialyzed individuals on additional immunosuppressive medication (yellow circles, n=10). Correlation was calculated using Spearman’s coefficient.

## 4. Discussion

In order to control the ongoing COVID-19 pandemic, efficient vaccination to create herd immunity without the infection-induced mortality will be key. We initiated this study to further increase information, including neutralization and response to variants of concern, on vaccination-induced immune responses in at risk immunocompromised populations such as renal dialysis patients. Similar to other groups, we found robust efficacy of the vaccine within dialyzed patients (9, 13), confirming that our results are comparable to other studies. Overall, 95% of our dialysis patients showed a humoral immune response to vaccination, higher than what other studies with similar time points had found (10-12, 14, 25). We can only speculate about potential reasons such as difference in renal replacement therapies, composition of the patient cohort or co-morbidities. It should be noted that titers were significantly reduced in dialysis patients compared to control individuals, which could result in reduced vaccine efficacy within this group. We also observed differences in humoral IgG and cellular T-cell response in our dialysis group, with four non-humoral responders and 23 non-T-cell responders, respectively. Overall, it is apparent in our data that in both groups vaccination response was skewed towards secretory immunity. This is in line with exploratory studies using BNT161b1 and other mRNA vaccines, which induce a B-cell response peak around two weeks after the boosting dose to then decline before reaching a memory plateau phase (26, 27). Future studies will be needed to determine the longevity and relative contribution of both T- and B-cell responses towards vaccination-induced protection. Interestingly, we found no significant differences in vaccination-induced IgA levels in both saliva and plasma between our study groups. Although several studies reported lower protective IgG titers over time following hepatitis B, influenza A or SARS-CoV-2 vaccinations, IgA levels were not analyzed here (9-17). Interestingly, a monoclonal IgA antibody capable of recognizing both the SARS-CoV-1 and SARS-CoV-2 spike proteins and blocking ACE2 receptor interaction combined with an increased neutralization ability over its IgG equivalent has been described (28). .Some studies even report a higher neutralizing capacity of purified serum IgA monomers from early convalescent sera compared to IgG and increased saliva IgA titers and neutralization versus IgG in recovered hospitalized COVID-19 patients (29). We identified a clearly reduced neutralizing capacity towards all VoC RBDs tested in our dialyzed individuals compared to controls. Taking into account that SARS-CoV-2 infections were increased in vaccinated dialysis patients compared to vaccinated control individuals (11), further monitoring is urgently needed to determine if vaccine-induced protection prevents infection with increasingly circulating and diverse SARS-CoV-2 mutant strains or if additional protection measures still need to be put in place throughout therapy session despite a completed vaccination scheme.

Individuals with kidney failure are at increased risk of infections and malignancies and the uremic milieu may trigger a chronic inflammatory state, which promotes T-cell exhaustion and suppression of IFN γ production (30, 31). Indeed, patients with end-stage kidney disease (ESKD) had significantly higher serum levels of cytokines such as IFNγ, TNFα, IL-8, and CCL-2 compared to healthy controls. After mitogen stimulation, both CD4^+^ and CD8^+^ T-cells in ESKD group demonstrated a pro-inflammatory phenotype, more exhausted and anergic CD4^+^ and CD8^+^ T-cells and a reduced frequency of follicular helper T-cells, which are important for humoral immunity (32). In the light of these immunological changes, our results about diminished SARS-CoV-2 specific T-cell responses and increased proinflammatory cytokine release may account in part for impaired vaccine-induced IgG responses in these patients.

Our study has several limitations. While we have a reasonable sample size (81 dialysis patients), which is similar or even larger compared to several other studies (12, 13, 25), our control group is not age- and gender-matched. In addition, we evaluated only one of the currently approved SARS-CoV-2 vaccines with samples from a single dialysis center and did not perform in-depth immune phenotyping or assessment of SARS-CoV-2 responsive T-cell frequencies. We also lack paired saliva and plasma samples pre- and post first dose to characterize B- and T-cell response kinetics or assess potential cross-reactivity of endemic CoV antibodies in immunocompromised individuals across the dosing scheme. However, all of our samples were collected at the same time and following an identical dosing regimen, allowing us to make a direct comparison between our two groups of interest (dialyzed versus non-dialyzed). Additionally, the lack of previously infected samples within our study groups, means we are studying only the vaccine-induced response.

Taken together, we provide robust evidence that a completed two-dose regimen of BNT162b2 elicits both antibody and T-cell responses in patients on maintenance dialysis towards the SARS-CoV-2 B.1 isolate. Future studies are needed to assess the lifespan and long-term kinetics of the vaccination response. As neutralization is reduced in dialyzed patients towards all VoCs examined, our data also highlights the need to monitor if infection with SARS-CoV-2 variants of concern occur more frequently in this vulnerable population compared to vaccinated healthy individuals.

## Supporting information

Supplementary Material Strengert et al.

## Data Availability

Data relating to the findings of this study are available from the corresponding authors upon request.

## Contributors

AD-J, GMNB, MSN, GL and MS conceived the study. MB, AD, MS, AD-J, GMNB, AC, NSM and MVS designed the experiments. NSM, MS, GMNB, AD-J, and GK procured funding. GMR, JG, JJ and MVS performed experiments. KL, AB, EW, GL, AC, and GMNB collected samples or organized their collection. PDK, BT and UR produced and designed recombinant assay proteins. MB, AD, MS, GMR, MVS and AC performed data collection and analysis. MB generated the figures. MS wrote the first draft of the manuscript with input from MB, NSM, GMNB and AD. All authors critically reviewed the final manuscript.

## Acknowledgments

We sincerely thank all patients for their contribution and willingness to participate in this study. We also thank the clinical staff for their additional efforts to make this study possible.

