## Supplementary Material Strengert et al. for "Cellular and humoral immunogenicity of a SARS-CoV-2 mRNA vaccine in patients on hemodialysis"

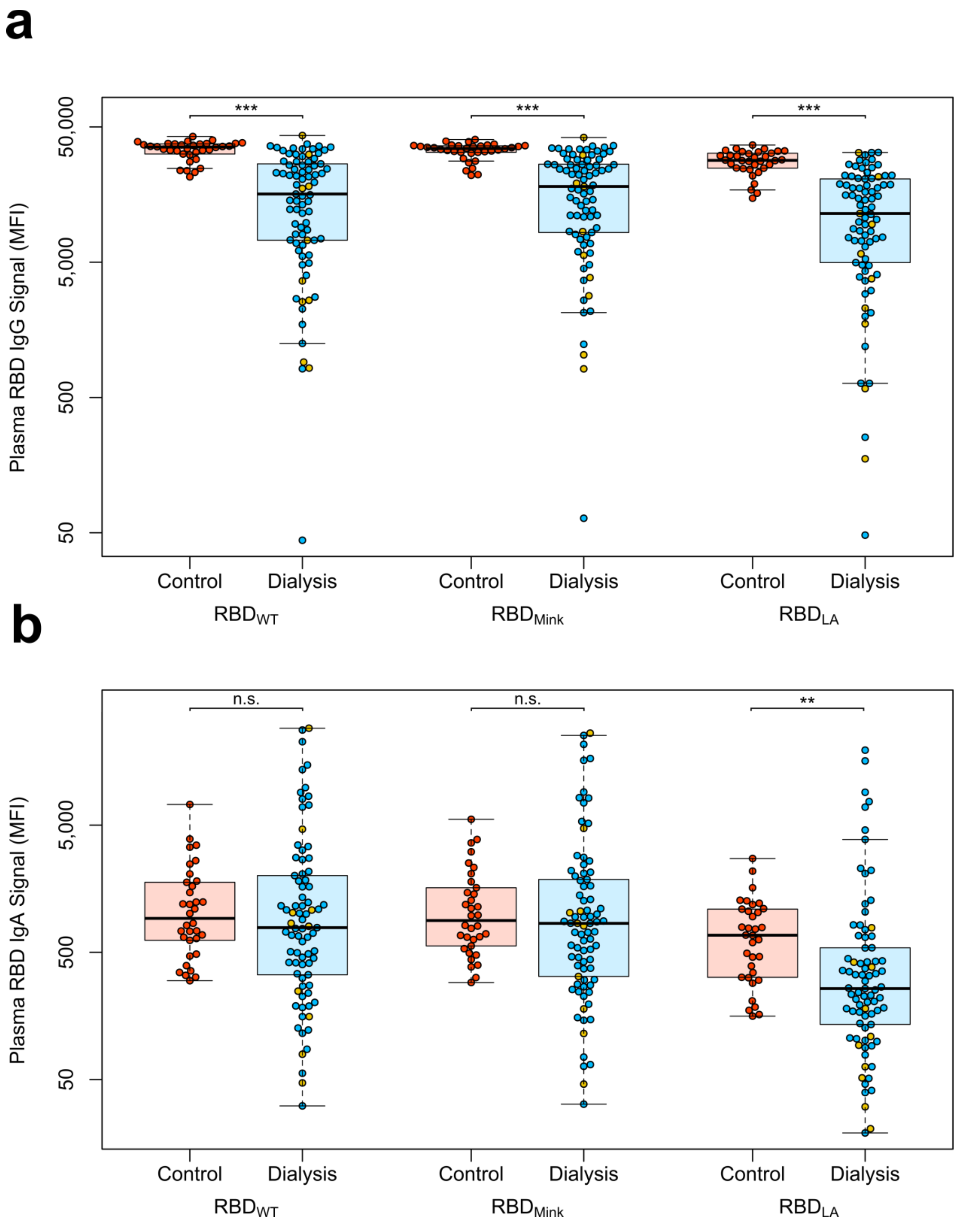


**Fig. S1. Binding capacity of antibodies from plasma after vaccination with Pfizer BNT162b2 for RBDs of VoC B.1.429 (LA) and Cluster 5 (Mink).**

Plasma IgG (a) and IgA response (b ) towards the indicated SARS-CoV-2 VoC cluster 5 (Mink) and B.1.429 (LA) from controls (red circles, n=34), individuals on maintenance dialysis (blue circles, n=71) and hemodialyzed individuals on immunosuppressive medication (yellow circles, n=10) 21 days post second vaccination was measured using MULTICOV-AB. Data is displayed as median fluorescence intensity (MFI). Boxes represent the median, 25th and 75th percentiles, whiskers show the largest and smallest non-outlier values. Outliers were determined by 1·5 times IQR. Statistical significance was calculated by Mann-Whitney-U (two-sided). Significance was defined as *<0·01, **<0·001, ***<0·0001 or n.s.>0·01. Binding towards wild-type RBD (B.1) is again displayed for clarity and comparison.

.

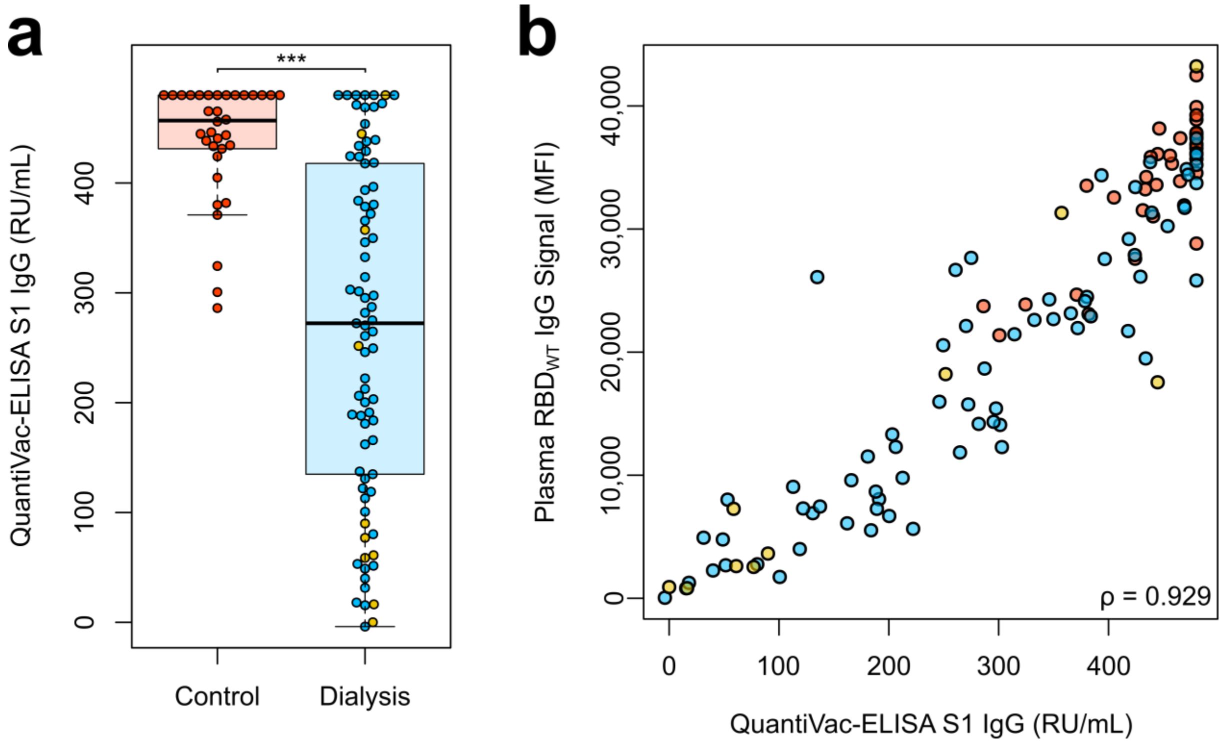


**Fig. S2. Quantitative IgG titers in plasma after vaccination with Pfizer BNT162b2.**
(a) Spike subdomain 1 (S1)-plasma IgG from controls (red circles, n=34), individuals on maintenance dialysis (blue circles, n=71) and hemodialyzed individuals on immunosuppressive medication (yellow circles, n=10) 21 days post second vaccination were analyzed using the QuantiVac-ELISA from Euroimmun (RU/mL). Samples that were classified as above upper or below the lower limit of detection of the assay are shown at the respective limit. Boxes represent the median, 25th and 75th percentiles, whiskers show the largest and smallest non-outlier values. Outliers were determined by 1·5 times IQR. Statistical significance was calculated by Mann-Whitney-U (two-sided). Significance was defined as *<0·01, **<0·001, ***<0·0001 or n.s.>0·01. (b) Correlation of MULTICOV-AB wild-type RBD B.1-IgG and Euroimmun Quantivac Spike S1-IgG is shown across the study cohort. Correlation was calculated using Spearman´s coefficient.

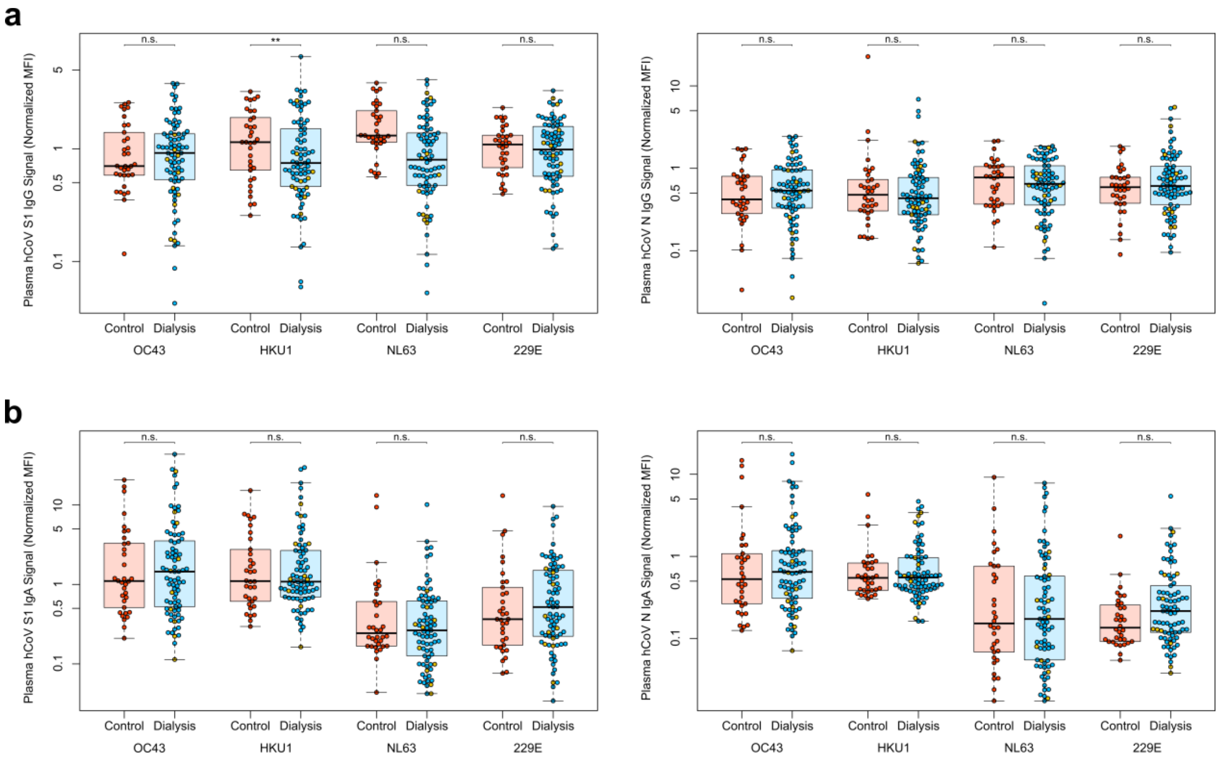


**Fig. S3. Humoral immune response towards endemic coronaviruses in the vaccination cohort.**
Plasma IgG (a) or IgA (b) levels towards S1 (left panel) or nucleocapsid (N, right panel) antigens were measured using MULTICOV-AB. Data for controls (red circles, n=34), individuals on maintenance dialysis (blue circles, n=71) and hemodialyzed individuals on immunosuppressive medication (yellow circles, n=10) is shown as normalized MFI using quality control samples for IgG and IgA, respectively. Boxes represent the median, 25th and 75th percentiles, whiskers show the largest and smallest non-outlier values. Outliers were determined by 1·5 times IQR. Statistical significance was calculated by Mann-Whitney-U (two-sided). Significance was defined as *<0·01, **<0·001, ***<0·0001 or n.s.>0·01.

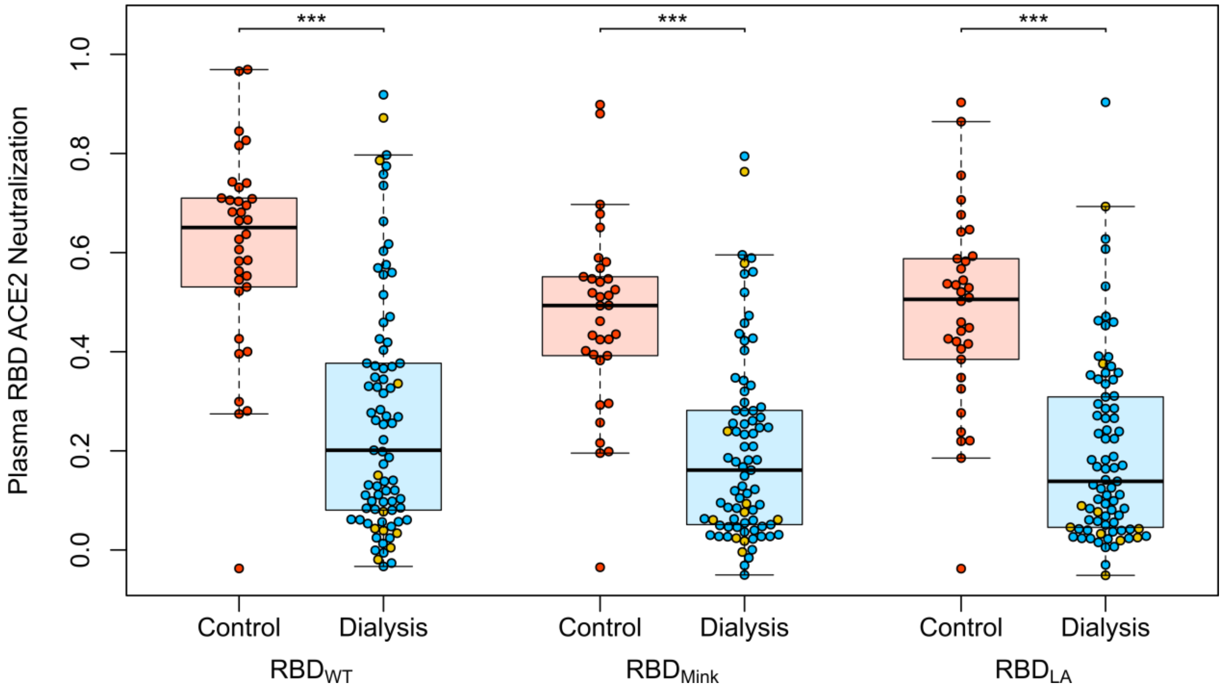


**Fig. S4. Neutralizing capacity of plasma IgG after vaccination with Pfizer BNT162b2 for RBDs of VoC B1.429 (LA) and Cluster 5 (Mink).**

Neutralizing capacity of plasma IgG towards the indicated SARS-CoV-2 variants of concern cluster 5 (Mink) and B.1.429 (LA) from controls (red circles, n=34), individuals on maintenance dialysis (blue circles, n=71) and hemodialyzed individuals on immunosuppressive medication (yellow circles, n=10) 21 days post second vaccination was measured using an ACE2-RBD competition assay. Neutralization capacity is displayed as ratio where 1 indicates maximum neutralization and 0 no neutralization. Boxes represent the median, 25th and 75th percentiles, whiskers show the largest and smallest non-outlier values. Outliers were determined by 1·5 times IQR. Statistical significance was calculated by Mann-Whitney-U (two-sided). Significance was defined as *<0·01, **<0·001, ***<0·0001 or n.s.>0·01. Neutralization towards the wild-type RBD (B.1) is again displayed for clarity and comparison.

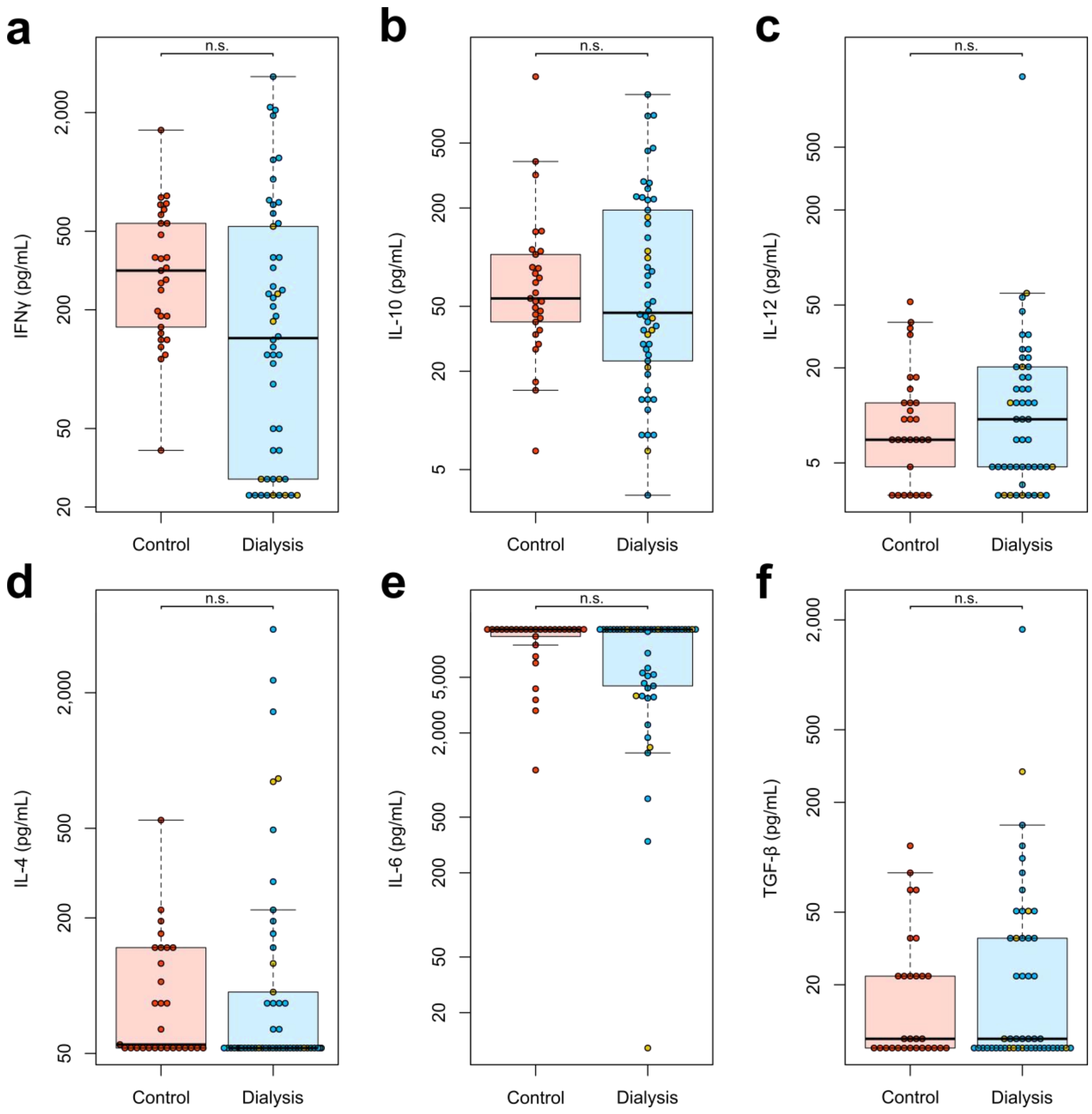


**Fig. S5. Cytokine responses from PBMCs after *ex vivo* stimulation using a Spike S1-specific peptide pool.** Whole blood from vaccinated controls (red circles, n=29), individuals on maintenance dialysis (blue circles, n=40) and hemodialyzed individuals on immunosuppressive medication (yellow circles, n=10) 21 days post second vaccination was *ex vivo* stimulated using a SARS-CoV-2 spike S1-specific peptide pool. Supernatant fractions were analyzed by bead-based multiplex-cytokine assay (pg/mL) for IFNγ (a), IL-10 (b), IL-12 (c), IL-4 (d), IL-6 (e) and TGF-β (f). Samples that were classified as above upper or below the lower limit of detection of the assay are shown at the respective limit. Boxes represent the median, 25th and 75th percentiles, whiskers show the largest and smallest non-outlier values. Outliers were determined by 1·5 times IQR. Statistical significance was calculated by Mann-Whitney-U (two-sided). Significance was defined as *<0·01, **<0·001, ***<0·0001 or n.s.>0·01.

**Table S1. MULTICOV-AB antigen panel**

| Virus | Antigen | Manufacturer | Product number |
|---|---|---|---|
| SARS-CoV-2 | Spike Trimer | NMI | - |
| SARS-CoV-2 | RBD B.1 (wild-type) | NMI | - |
| SARS-CoV-2 | Nucleocapsid | Aalto | 6404-b |
| SARS-CoV-2 | RBD B.351 (South Africa) | NMI | |
| SARS-CoV-2 | RBD B.1.1.7 (UK) | NMI | |
| SARS-CoV-2 | RBD Cluster 5 (Mink) | NMI | |
| SARS-CoV-2 | RBD B.4.129 (LA) | NMI | |
| hCoV-OC43 | S1 domain | NMI | - |
| hCoV-OC43 | Nucleocapsid | NMI | - |
| hCoV-HKU1 | S1 domain | NMI | - |
| hCoV-HKU1 | Nucleocapsid | NMI | - |
| hCoV-NL63 | S1 domain | NMI | - |
| hCoV-NL63 | Nucleocapsid | NMI | - |
| hCoV-229E | S1 domain | NMI | - |
| hCoV-229E | Nucleocapsid | NMI | - |

**Table S2.** Causative kidney disease for dialysis – ANCA - Anti-Neutrophilic Cytoplasmic Autoantibody.

| Characteristics | Hemodialysis group (n=81) |
|---|---|
| Diagnosis (n, %) | |
| Autosomal dominant polycystic kidney disease | 11 (13·58) |
| Chronic glomerulonephritis | 6 (7·41) |
| Diabetic nephropathy | 13 (16·05) |
| Focal segmental glomerulosclerosis | 5 (6·17) |
| IgA nephropathy | 9 (11·11) |
| Interstitial nephropathy | 6 (7·41) |
| Nephrosclerosis | 18 (22·22) |
| Acute toxic tubular epithelial damage syndrome | 1 (1·23) |
| Primary amyloidosis | 1 (1·23) |
| ANCA-associated vasculitis | 1 (1·23) |
| Cardiorenal syndrome | 2 (2·47) |
| Medullary cystic kidney disease | 1 (1·23) |
| Membranous glomerulonephritis | 1 (1·23) |
| Kidney dysplasia | 1 (1·23) |
| Obstructive nephropathy | 1 (1·23) |
| Reflux nephropathy | 1 (1·23) |
| Septic organfailure | 1 (1·23) |
| Cystic kidney disease | 1 (1·23) |
| Cyclosporin intoxication | 1 (1·23) |

**Table S3.** Medication of vaccinated study participants.

| Characteristics | Non-dialysis control group (n=34) | Hemodialysis group (n=81) |
|---|---|---|
| Medication (n, %) | | |
| Angiotensin-converting enzyme inhibitors | 2 (5·88) | 25 (30·86) |
| Statins | 0 (0) | 47 (58·02) |
| Angiotensin II Receptor Blockers | 9 (26·47) | 26 (32·10) |
| Vitamin D Supplements | 16 (47·06) | 80 (98·77) |
| Immunosuppressants | | |
| Prednisolon | 0 | 6 (7·41) |
| Prednisolon, Tacrolimus | 0 | 2 (2·47) |
| Prednisolon, Tacrolimus, Mycophenolatmofetil | 0 | 1 (1·23) |
| Hydrocortisone | 0 | 1 (1·23) |
| Budesonide | 0 | 1 (1·23) |
